# Inclusion Criteria for Extracorporeal Membrane Oxygenation (ECMO) in Patients with Acute Respiratory Distress Syndrome (ARDS) Due to COVID-19: A Systematic Review

**DOI:** 10.1101/2023.07.01.23291847

**Authors:** Panagiota Triantafyllaki, Marios Charalampopoulos, Christina-Athanasia Sampani, Christos Triantafyllou, Dimitrios Papageorgiou

**Author notes:** **Corresponding author:** Christos Triantafyllou, Papadiamantopoulou 123, Goudi Athens, PC 11527. On Behalf: ICU Follow-Up Care Lab, Department of Nursing, University of West Attica, Athens, Greece.

## Abstract

**Introduction:** At the end of 2019, in the city of Wuhan, China, a virus of the family of coronaviruses first appeared, mainly affecting the respiratory system, which was called SARS-COV-2 and causes COVID-19. Although in most patients, it occurs with mild symptomatology, however, a significant percentage (15-30%) will develop acute respiratory distress syndrome (ARDS) with increased chances of intubation and mechanical ventilation. In special cases of severe disease, where the oxygenation of the patient is not improved by the use of the ventilator, extracorporeal membrane oxygenation (ECMO) can be applied, a technique that has been used in previous pandemics that affected the respiratory system.

**Aim:** To investigate the evidence of the appliance of the ECMO, based on international literature, of the extracorporeal membrane oxygenator in patients with severe respiratory failure due to Covid-19 disease.

**Method:** Articles were searched on the international bases of scientific studies PubMed, Cochrane Library, and Google Scholar. This review was carried out using meta-analysis and international guidelines.

**Results:** Four articles were included where there was an agreement on the basic characteristics of patients, which can be considered as selection criteria. The primary criteria indicate the age, where the patient must be under 65 years old, and the body mass index (BMI) should be below 40. In addition, it is very important that there is no serious underlying pathology such as multi-organ failure syndrome. Also, the mechanical ventilation should not exceed seven (7) days until the placement of the ECMO, while all the other therapeutic methods, such as the prone position, neuromuscular blockers, and the appropriate positive end-expiratory pressure of the airways (Positive end-expiratory pressure - PEEP) should be already applied.

**Conclusions:** The application of ECMO is widely used as a treatment for patients with severe COVID-19 disease. However, in order to have the best therapeutic results while reducing hospitalization costs, it is necessary to follow the guidelines regarding the selection of patients who will benefit substantially.

## Introduction

At the end of 2019, in the city of Wuhan, China, a virus of the coronavirus family, mainly affecting the respiratory system, which was named SARS-COV-2 and causes the disease COVID-19, appeared for the first time. Although most patients present with mild symptoms, a significant proportion (15-30%) will develop severe respiratory distress (ARDS) with increased chances of intubation and mechanical respiratory support.^1,2,3^

Acute respiratory distress syndrome (ARDS) is an acute inflammatory pulmonary process that leads to protein-rich, non-hydrostatic pulmonary edema, unwanted hypoxemia, and lung stiffness. As a result, there is an inability to eliminate carbon dioxide and a disturbance in gas exchange. Clinically, it is characterized by acute respiratory failure, bilateral infiltrates on the chest X-ray, and hypoxemia with the PaO2/FiO2mmHg index evaluation. However, no evidence of left atrial hypertension or pulmonary capillary pressure should be observed. ^4,5^

In special cases of severe illness, where the patient’s oxygenation does not improve with the ventilator, ECMO can be applied, a technique that has also been applied in previous pandemics that affected the respiratory system. ^5,6,7^

Depending on the pathological entity being treated, ECMO can be used to support the respiratory and/or cardiovascular system. In addition, regarding respiratory support through an extracorporeal circuit, which includes large-diameter catheters, the blood is led outside the body to the oxygenator membrane where gas exchange takes place, Veno-venous ECMO is a technique that two central venous vessels are catheterized, and the blood flow is being recycled ^6,7^.

Accordingly, in patients with severe heart failure (with or without respiratory distress), a veno-arterial connection of the catheters is performed, where the blood from the central vein is led to the oxygenator and returns to the central artery (veno-arterial ECMO).^6,7^

Despite the potentially beneficial effect of ECMO, this technique cannot be universally applied to all patients admitted with ARDS due to COVID-19 in the ICU. To ensure the best possible results from the use of ECMO, it is necessary to apply specific selection criteria for patients who will be supported extracorporeally and to distinguish those patients who will not benefit and, therefore, must be excluded.^7^

The purpose of this study is to gather the information that exists so far, based on the international literature, regarding the indications for the introduction of the external membrane oxygenator in patients with severe respiratory failure due to the covid-19 disease. The individual objectives of the study it is to find the qualitative characteristics of the patients aggregated, which are likely to be associated with the best possible prognosis, based on clinical findings such as the improvement of the aerometric image of the hospitalized patient. Finally, among the individual objectives, the reference to the absolute contraindications to the application of ECMO is included since the condition of the patients is not amenable to improvement.

## Method

Articles were searched in international databases of scientific studies, PubMed, Cochrane Library, and Google Scholar. This review was performed using systematic reviews, meta-analyses, and international guidelines. The following terms were used as keywords: “ECMO”, “ECMO criteria”, “ECMO guidelines”, “ARDS”, “COVID-19 treatment”, and “ICU”. In addition, a combination of the above terms was used for more targeted article findings. Criteria for inclusion and exclusion of the studies under evaluation were set. The following were used as inclusion criteria:

- Studies carried out in the last ten years (2012-2022),
- Articles with free access.
- Studies conducted in human population.
- Studies conducted in an adult population.
- Studies involving the VV ECMO method.

The following were introduced as exclusion criteria:

- Studies involving animals.
- Studies published in a language other than English.

The process of selecting and excluding articles is captured in **Figure 1**.

**Figure 1:**
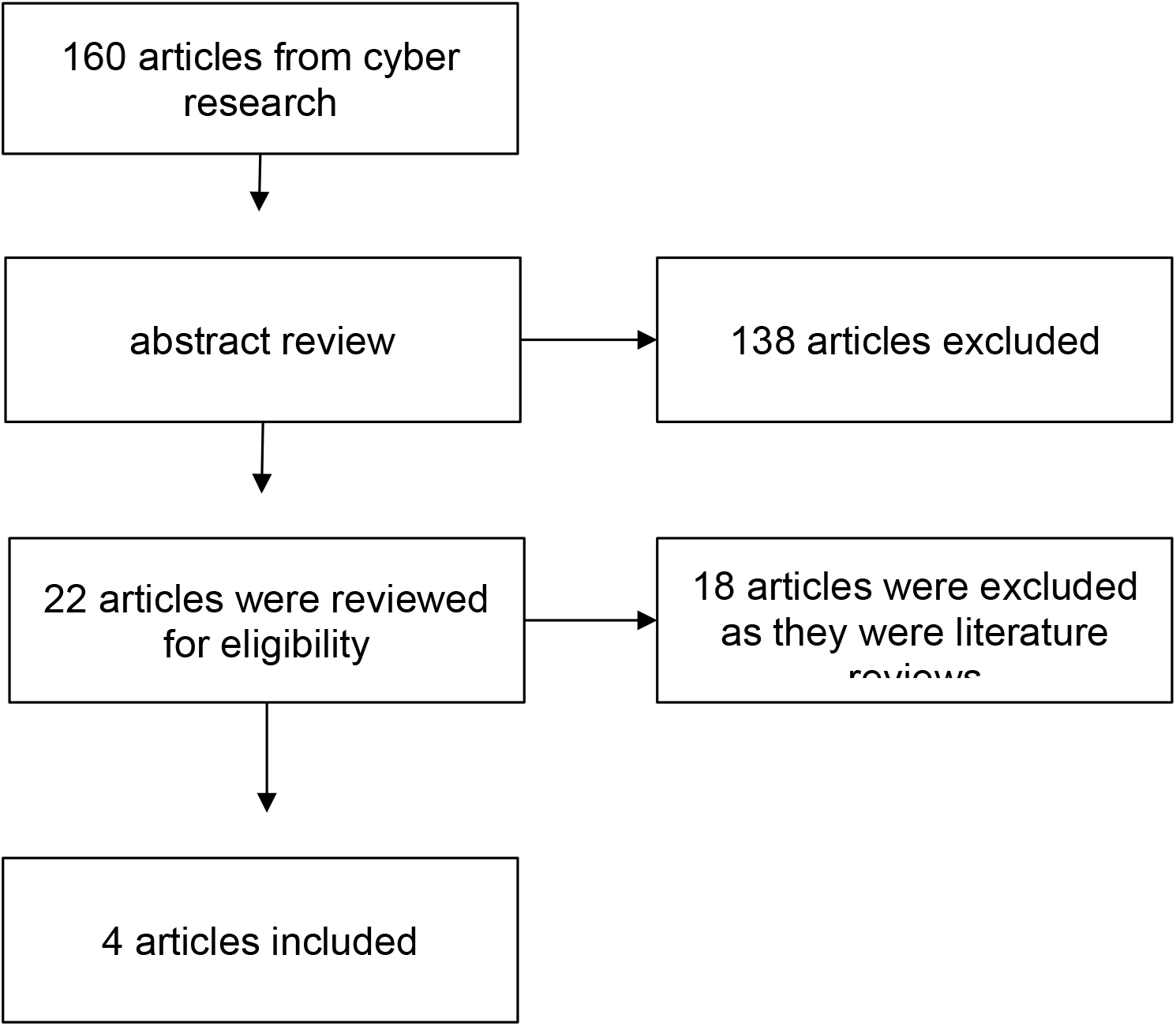
Flow chart of the systematic review.

Finally, four studies were included. These studies came from Boston (one study), Minnesota (one study), Geneva (one study), and Amsterdam (one study). No quantitative synthesis of results was performed, but only a systematic review of studies. Also, no assessment of the quality of the studies based on specific tools was attempted because the purpose of the specific research was descriptive.

## Results

In the present systematic review, an attempt was made to present the basic criteria for the introduction of ECMO in patients with a severe form of ARDS COVID-19 disease. The severe respiratory failure created as a pathophysiological progression of the disease does not allow the usual therapeutic procedures to cause a satisfactory improvement of the patient’s clinical picture, such as, for example, protective mechanical ventilation and the use of sedation in combination with neuromuscular blockers. Thus, it is necessary, in a short period of time from the start of the patient’s mechanical respiration (up to 7 days), and if there is no clinical improvement of the respiratory function, to apply ECMO to improve his oxygenation further. However, for ECMO to be applied and for the patient to effectively benefit from this mechanism, it is necessary to ensure that the person does not succumb to contraindications such as multi-organ failure, advanced age, some chronic end-stage disease, and others. The results of the present systematic review are presented in **Table 1**.

**Table 1.**
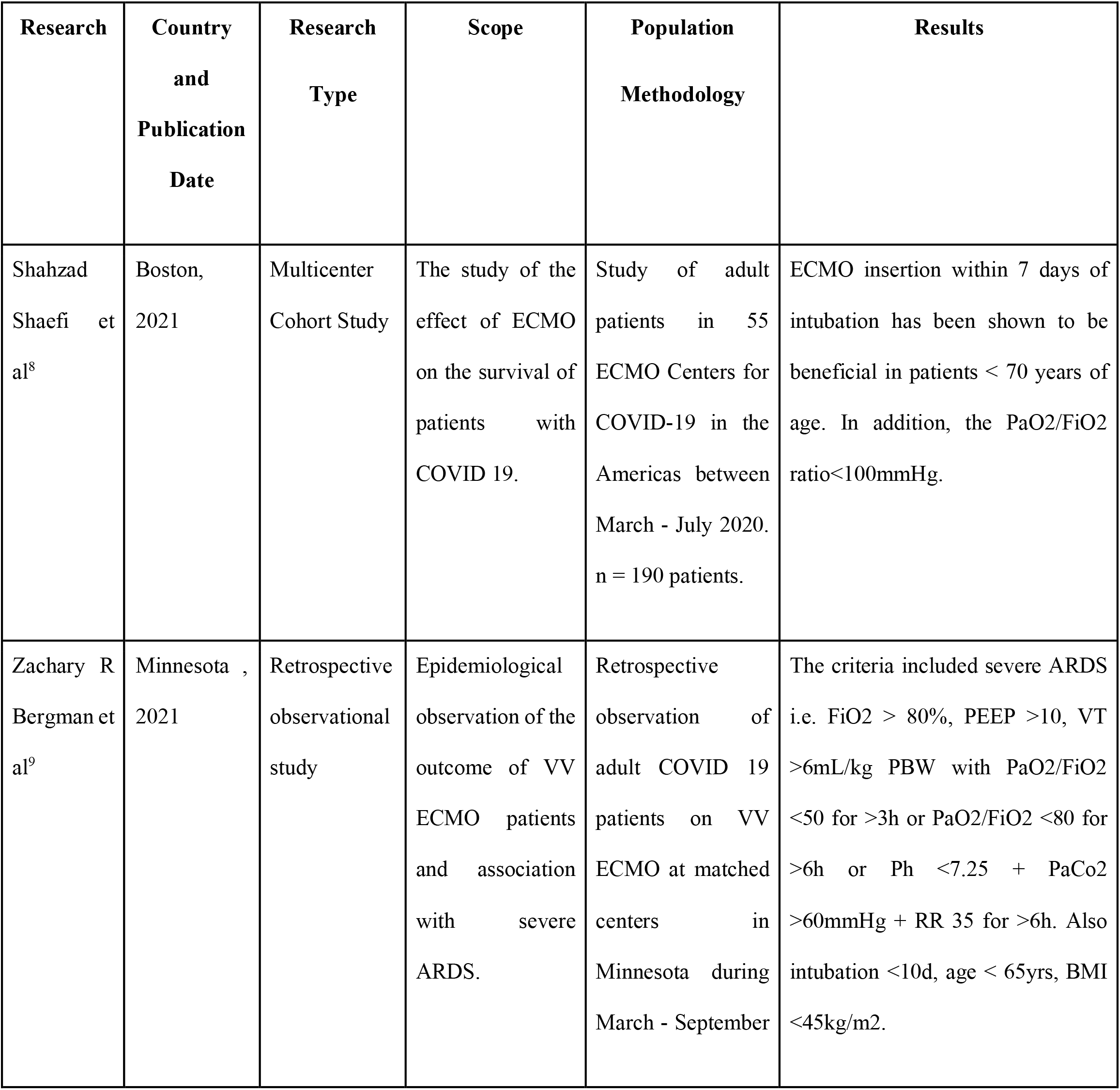
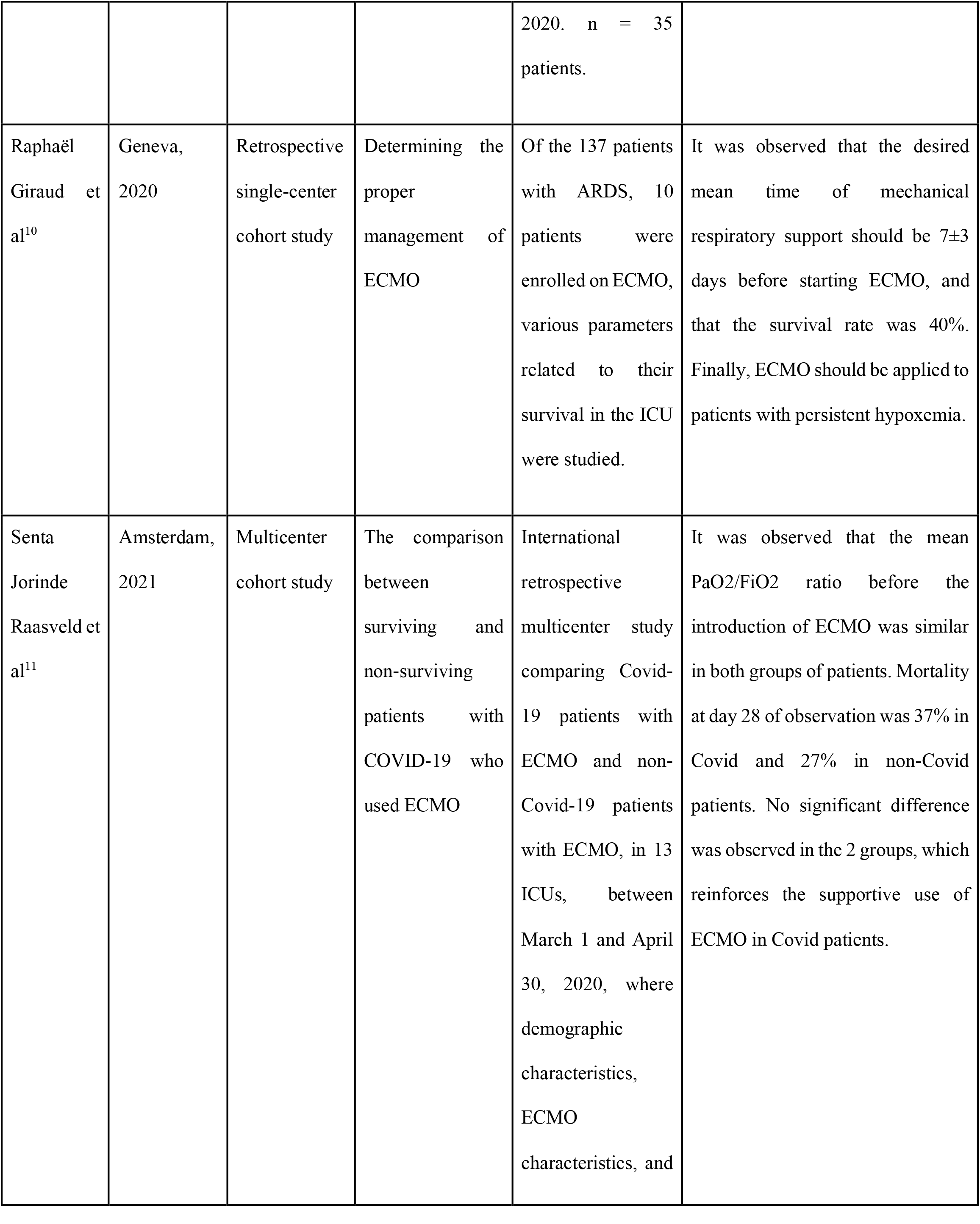
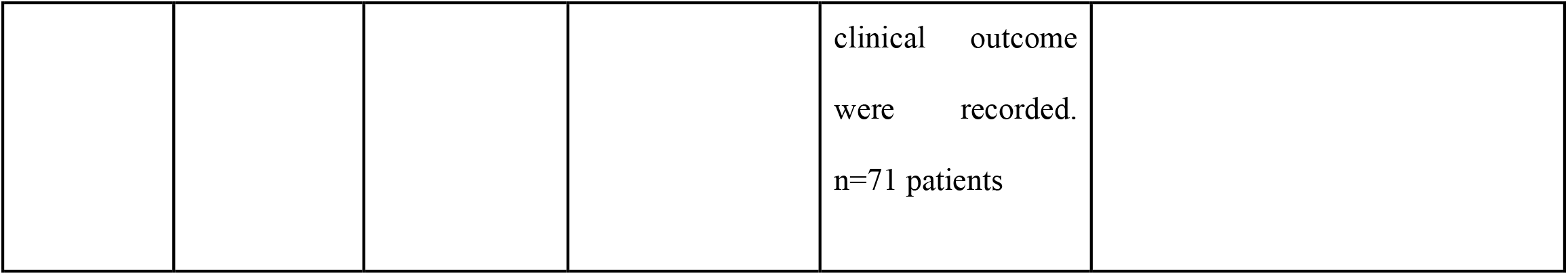
Summary of the studies in the systematic review

### Shahzad Shaefi et al (2021)^8^

The study by Shahzad Shaefi et al., carried out in 55 Hospitals - Treatment Centers for COVID-19 using ECMO, enrolled a total of 190 ICU patients from March to July 2020. These patients were compared with those who survived or died within 60 days in order to derive results regarding the use of ECMO. In the ECMO admission selection criteria, based on the comparisons, seven (7) days after intubation were set as the desired upper limit of application of the membrane oxygenator as there were more chances of survival to a statistically significant degree. Of all patients, 83% were placed on ECMO within seven days, with an average of 3 days in the ICU. Also, a PaO2/FiO2 ratio <100mmHg while under mechanical ventilation was set as a criterion, as severe prolonged hypoxemia is associated with a poor prognosis. Finally, the upper age limit is set below 70 years. Of the patients supported with ECMO, 33.2% died.^8^

### Zachary R Bergman et al, (2021)^9^

In a retrospective observational study, which took place in specialized ECMO centers over a period of seven (7) months of the 1849 cases of COVID-19 admitted to the ICU, 35 were placed on ECMO VV support, and of this percentage, 26 patients survived (74%). The median day of admission from the day of intubation to the day of ECMO admission was day 3. Admission criteria included severe ARDS, i.e., FiO 2 > 80%, PEEP >10, VT >6mL/kg PBW, PaO2/FiO2<50 for more than 3 hours, or PaO2/FiO2<80 for more than 6 hours or Ph <7.25 and PaCo2 >60mmHg and respiratory rate 35 for more than 6 hours. Also, the patient must be intubated for less than ten days with an age of less than 65 years and finally, a body mass index (BMI) <45kg/m2.^9^

### Raphaël Giraud et al, (2020)^10^

In a single-center retrospective cohort study conducted in Switzerland under the auspices of the Swiss Society of Intensive Care, an algorithm was created for the appropriate application of ECMO in patients with Covid-19. Ten of the 137 ARDS cases admitted to the unit were selected to receive ECMO. Risk factors and comorbidities were recorded in these patients before they became infected with COVID. Additionally, on ICU admission and before ECMO placement, SAPPS II was measured, and a broader comparison of surviving and non-surviving ECMO patients was made. In the results, survivors had a significantly higher PH value (7.48 ± 0.03 vs. 7.32 ± 0.14, p=0.019) and a shorter intubation interval before receiving ECMO (91 ± 58 h vs. 208 ± 34 h, p = 0.01). No other comparisons revealed any statistically significant differences between groups.^10^

### Senta Jorinde Raasveld et al, (2021)^11^

In an international multicenter cohort study conducted among 13 Intensive Care Units between March 1 and April 30, 2020, a comparison was made between Covid-19 patients on ECMO and non-Covid-19 patients on ECMO. Among them, demographic characteristics, ECMO parameters, and clinical outcomes were compared. In the results, regarding ECMO admission criteria, it should be the last choice of supportive treatment in patients with acute respiratory distress syndrome and persistent hypoxemia. For this reason, all therapeutic agents, such as the prone position and neuromuscular blockers, should first be exhausted. Finally, applying to patients under 65 years of age 11 is recommended.^11^

## Discussion

ECMO as a support technique has been applied in the past in other pandemic cases, such as H1N1, to support the respiratory system. However, in these cases, the need for ECMO application was limited due to the clearly fewer cases of severe disease. During the COVID-19 pandemic, patients requiring hospitalization in an Intensive Care Unit due to Acute Respiratory Distress comparatively much more, which fact implies an increased need for the application of this therapeutic technique.^1,5^

In the majority of the studies included in this review, an agreement was observed regarding the basic characteristics of the patients, which can be considered selection criteria. The primary criteria include age, where the patient must be under 65, and a body mass index (BMI) under 40. In addition, it is very important that there is no serious underlying disease, such as heart failure and cancer end stage, but also multiple organ failure syndrome. At the level of therapeutic support, if the patient is on mechanical respiratory support, it should not exceed seven (7) days until ECMO placement, as it was shown that otherwise, survival rates are significantly reduced.^8,9,10^

Also, it is important to ensure that in intubated patients, before being supported with ECMO, maximal muscle relaxation and sedation, prone position technique, and appropriate PEEP have already been applied without satisfactory improvement in their respiratory profile. Regarding the evaluation of the respiratory profile for the application of ECMO, the majority of studies used the PaO2/FiO2 ratio, which is below 60mmHg for more than six (6) hours or PaO2/FiO2 below 50mmHg for more than three (3) hours or Ph below 7.20 and PaCo2 above 80mmHg for less than six (6) hours as, the smaller the ratio, the greater the chances for a poor patient outcome.^11,12,13^

Lastly, the strong contraindications to ECMO placement are coagulation disorders, hemorrhagic stroke, advanced lung fibrosis, advanced age, and the patient’s increased need for vasoconstrictor support.^5,6^

## Conclusions

The application of ECMO is widely used as a treatment for severely ill COVID-19 patients. However, to achieve optimal outcomes and reduce hospital costs, following guidelines for selecting patients who will benefit substantially is critical. Therefore, it is considered necessary to create and implement guidelines based on which patients will be categorized according to whether or not they will benefit from the use of ECMO in order to ensure the best outcome for patients. However, the short period of study of the entire process in the context of the COVID-19 era requires further study on ECMO research centers to review and add additional criteria that will ensure the best outcome for the patient.

## Data Availability

All data produced in the present study are available upon reasonable request to the authors.

